# Remote olfactory assessment using the NIH Toolbox Odor Identification Test and the Brain Health Registry

**DOI:** 10.1101/2020.12.14.20248177

**Authors:** Cristina Jaén, Christopher Maute, Scott Mackin, Monica R. Camacho, Diana Truran, Rachel Nosheny, Michael W. Weiner, Pamela Dalton

## Abstract

**Objective:** Early identification of deficits in our ability to perceive odors is important as many normal (i.e., aging) and pathological (i.e., sinusitis, viral, neurodegeneration) processes can result in diminished olfactory function. However, unlike hearing and vision, olfactory function is rarely tested outside of a research laboratory. The purpose of this study is to determine the feasibility of remotely testing olfactory performance using the National Institutes of Health’s Toolbox odor identification test (NIH Toolbox Odor ID Test).

**Methods:** Participants were recruited using the Brain Health Registry (BHR), an online assessment platform which connects participants with researchers. Interested participants were mailed the NIH Toolbox Odor ID Test along with instructions on accessing a website to record their responses.

**Results:** Data obtained from subjects who performed the test at home was comparable to the normative data collected when the NIH Toolbox Odor ID Test was administered by a tester in a research setting. Age-dependent olfactory decline and gender-dependent sensitivity could be detected using the NIH Toolbox Odor ID Test remotely.

**Conclusions:** The NIH Toolbox Odor ID Test is a valid instrument to measure olfactory performance via self-administration at home. This approach can be useful for longitudinal studies or as a screening tool for studies that require testing the sense of smell.

## Introduction

Our sense of smell provides information about our environment and is vital for critical functions like: eating, detecting noxious odors, and social functioning; while impaired olfaction negatively affects quality of life thereby impacting on physical and mental well-being, as well as social relationships. Decreased or absent olfactory function (hyposmia or anosmia, respectively) is estimated to afflict 3–20% of the population [1]. Unfortunately, many individuals with olfactory dysfunction are not aware of their problem until tested. Our sense of smell diminishes naturally with age, beginning around the 5th decade of life [2]. Women generally outperform men in olfactory abilities [3]. Multiple factors can compromise smell function, including: allergies, bacterial and viral infections, head injuries, sino-nasal disease, and environmental exposures to toxins and pollutants [4]. Importantly, smell dysfunction is one of the hallmark “preclinical” signs of neurodegenerative diseases such as Alzheimers and Parkinsons [5-7]. A recent longitudinal study found that olfactory dysfunction is one of the strongest predictors of 5-year mortality in older adults [8] as well as a predictor of dementia [9]. Overall, olfactory function appears to be a good indicator of general health [10, 11].

Measurement of olfactory ability in the clinical or research setting typically consists of odor identification, odor discrimination, and odor detection threshold tasks [12]. Odor identification tests consist of the presentation of a supra-threshold concentration of an odor to a subject who must choose the appropriate odor name from several response options. The most common odor identification tests to date are the University of Pennsylvania Smell identification Test (UPSIT) [13], which is a scratch and sniff test with 40 microencapsulated odorants, and the Sniffin’ Sticks test, 16 odors that are presented in liquid form using felt-pens [14]. While these are excellent tests with normative data on many thousands of individuals, they are proprietary and thus the cost of usage in large studies can become prohibitive. As a part of the larger NIH Toolbox initiative, the NIH Toolbox Odor ID Test was created to be “off the shelf”, brief, inexpensive, and suitable for use during the whole lifespan [15]. The NIH Toolbox Odor ID Test is a “scratch-and-sniff” test that uses pictorial answers to identify odors and was originally designed with the intention of being administered with the participant and the tester in the same room. The adult version of the NIH Toolbox Odor ID Test consists of 9 odors that are microencapsulated and placed onto small individual cards. In this study we evaluated using this inexpensive test as a convenient and appropriate instrument to remotely assess the sense of smell. A major focus of the study was to determine the feasibility of remote unsupervised testing, recruiting individuals using the Brain Health Registry platform. We explored whether the mailing of the odor cards together would result in cross contamination of odors and whether unsupervised results obtained at home would replicate previous results from clinical settings.

### Methods

The study reported here was conducted in accordance with the Code of Ethics of the World Medical Association (Declaration of Helsinki) for experiments involving humans. It was approved by the University of Pennsylvania Institutional Review Board. All participants provided on line consent before starting the olfactory test.

### Participants

Participants were recruited through the BHR online registry. Participants aged 40 years or older were targeted for this study and were emailed study information by the BHR study team. 1,841 participants showed interest in participating in the smell study, and contacted the study team. About 76.75% (N = 1,413) responded by providing a mailing address, and 82.73% (N = 1,169) of those who received the odor identification test signed into the online platform, gave consent, and started the odor test. Three participants used incorrect codes, making it impossible to identify them. Three additional participants did not complete the test, claiming that they had problems using the website and/or had problems identifying the odors and voluntarily withdrew from the study. Six participants did the test twice due to problems with the odor cards or internet connection (their second response is the one chosen for analysis). For this study, we are using the responses of the 1,163 participants who completed the test and could be identified. Demographic information about our study participants is shown in table 1.

**Table 1.** Participants demographic information. The population ranged from 40 to 90 years old. A total of 1,163 participants completed this study.

| Gender |  | Race |  | Ethnicity |  | Age Distribution |  |
| --- | --- | --- | --- | --- | --- | --- | --- |
| Male | 287 | Caucasian | 1089 | Not Latino | 1117 | 40-49 | 38 |
| Female | 876 | Asian | 20 | Latino | 20 | 50-59 | 300 |
|  |  | Afric Am | 10 | No answer | 26 | 60-69 | 537 |
|  |  | Mixed | 27 |  |  | 70-79 | 266 |
|  |  | Pacific Islander | 1 |  |  | 80-90 | 22 |
|  |  | Native Am | 4 |  |  |  |  |
|  |  | Other | 9 |  |  |  |  |
|  |  | No answer | 3 |  |  |  |  |

Participants who contacted us and provided a mailing address received a letter that contained the 9 odor cards packed together in a small plastic bag along with instructions. To prevent possible cross-contamination, the original NIH Toolbox Odor ID Test cards were slightly modified in that each odor card was twice as tall as the originals so it could be folded top to bottom forming a cover, thus limiting odor cross-contamination. The provided instructions included: the link to the website that would record their responses, their study code, and how to sample the odors and make their online response.

The participants self-administered the test and recorded their answers on a secure online survey platform (www.surveymonkey.com). For each odor card, the online survey presented four possible odor identification responses as pictures with labels in English; only one was the target/correct odor identifier. To self-administer the stimulus, the participant was instructed to use a paper clip to scratch the micro-encapsulated odor patch on the card and smell the released odor. Responses were made by clicking on the chosen picture that corresponded to the odor card they were smelling, thus mimicking the way this test was originally performed. Feedback on the accuracy of their response was not provided. The NIH Toolbox Odor ID Test and a screenshot of the website are shown in figure 1.

**Figure 1.**
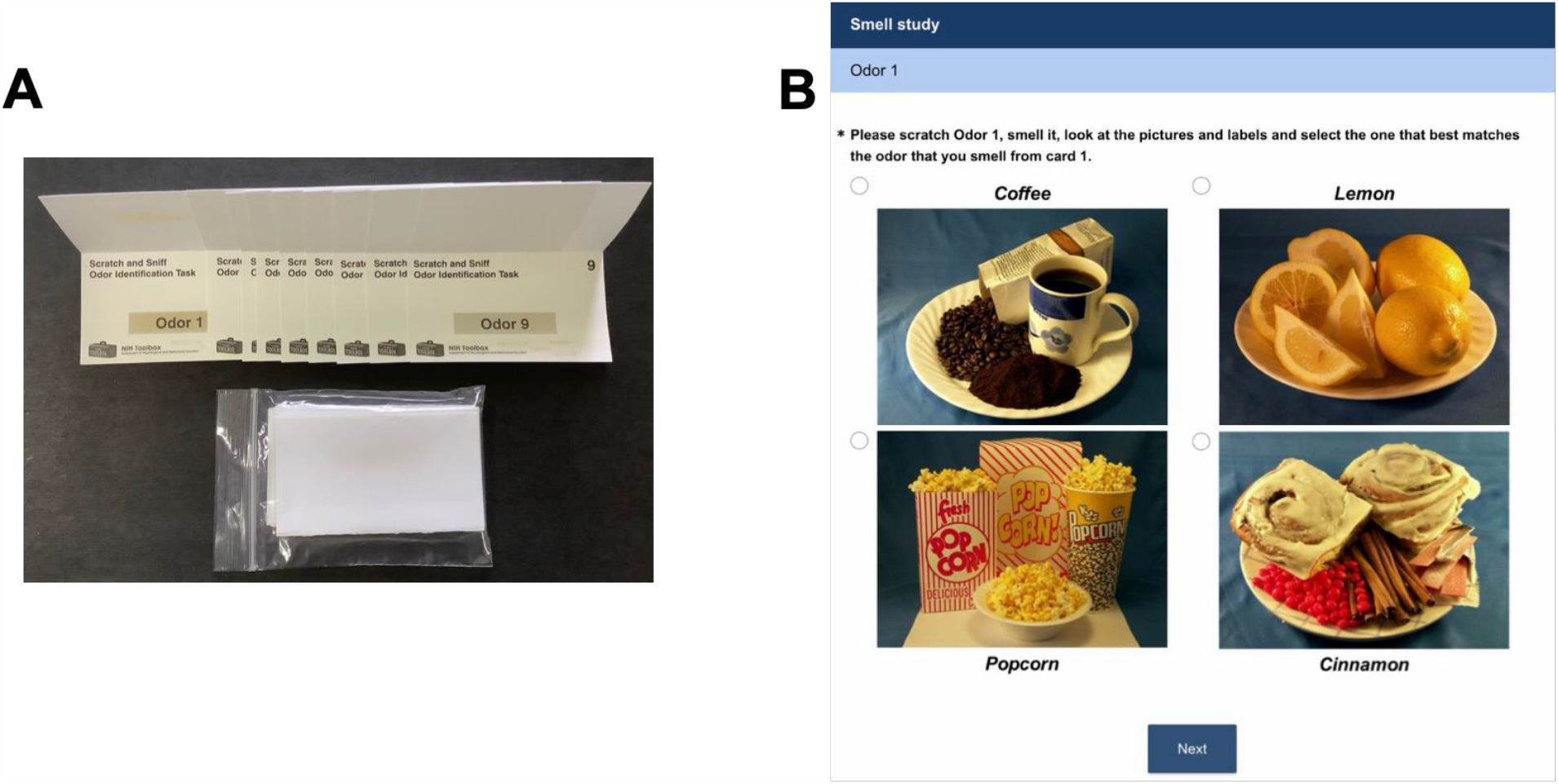
NIH Toolbox Oder ID Test. A) Modified Odor identification cards, each one has a cover to prevent odor cross_contamination. All nine folded odor identification cards were mailed together inside a small plastic bag B) Screenshot of the online platform showing the four possible answers for odor 1.

### Statistical Analysis

Data were analyzed used TIBCO Statistica software version 13.5. We used percentage of accurate odor response as olfactory accuracy as the dependent variable, and grouped participants into decades. For gender odor sensitivity analysis, we performed an ANCOVA using gender as factor and age as covariate and to analyze age-dependence olfactory decline we used a one-way ANOVA. Effect size for gender was calculated using Hedges’ g statistic.

## Results

As shown in figure 2, we found that sending the 9 odor id cards together did not cause odor cross-contamination. Odor integrity seemed to be maintained during the mailing process. Natural gas odor was the easiest to identify by 96% of the participants while chocolate was only correctly identified by 70% of the participants with 25% of the sample confusing it with a flower odor.

**Figure 2.**
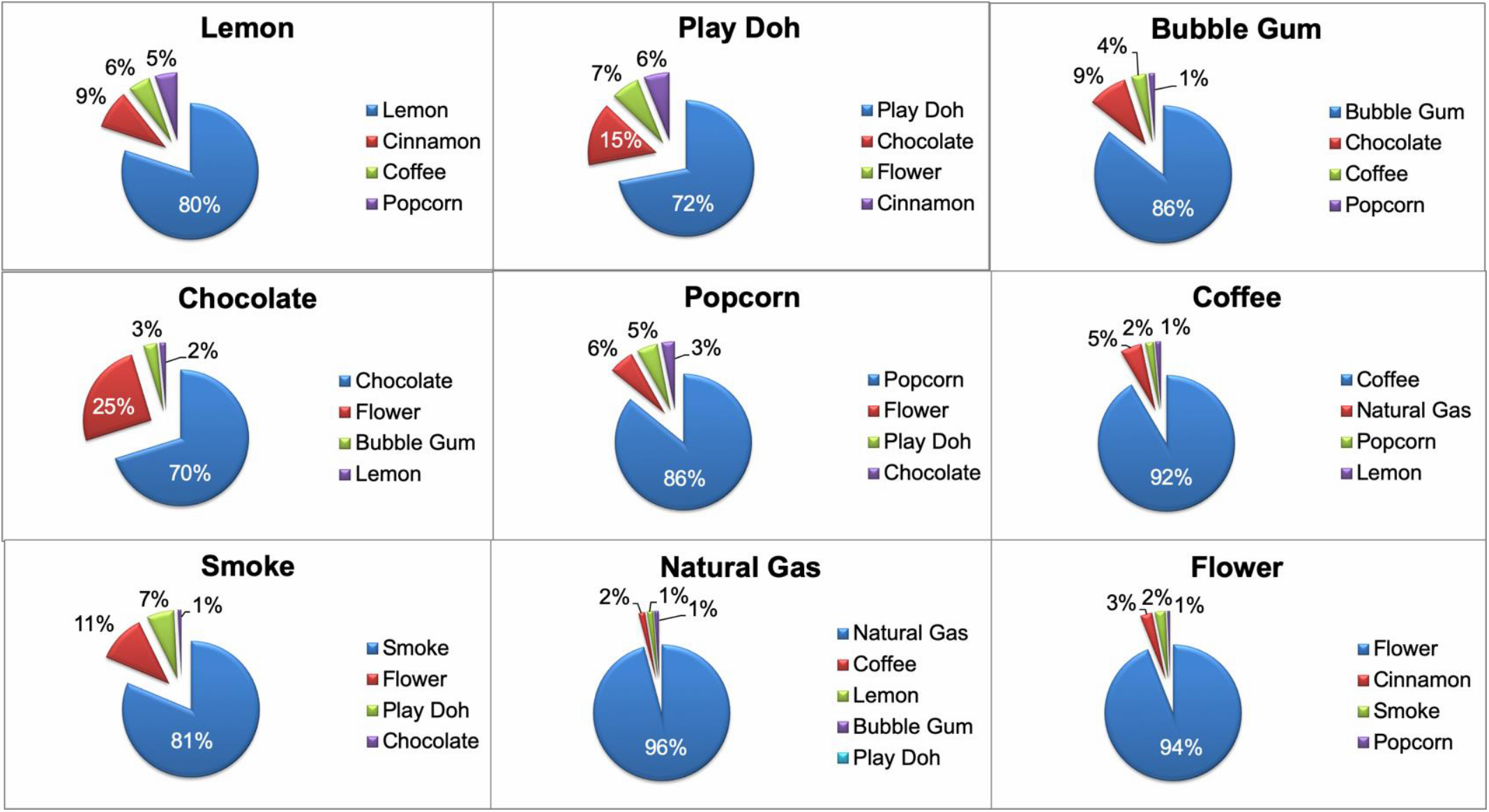
Odor identification attributed to each odor card. The correct response is at the header of each panel. Odor integrity was maintained after mailing the 9 odor ID cards together.

As shown in table 2, data obtained using the odor id cards either at home or in a testing place produced similar results. Some odors like natural gas, coffee, or flower seem to be easier to identify while others such as play Doh, chocolate and smoke seem to be more difficult to identify, and may depend more on personal experience with these odors.

**Table 2.**
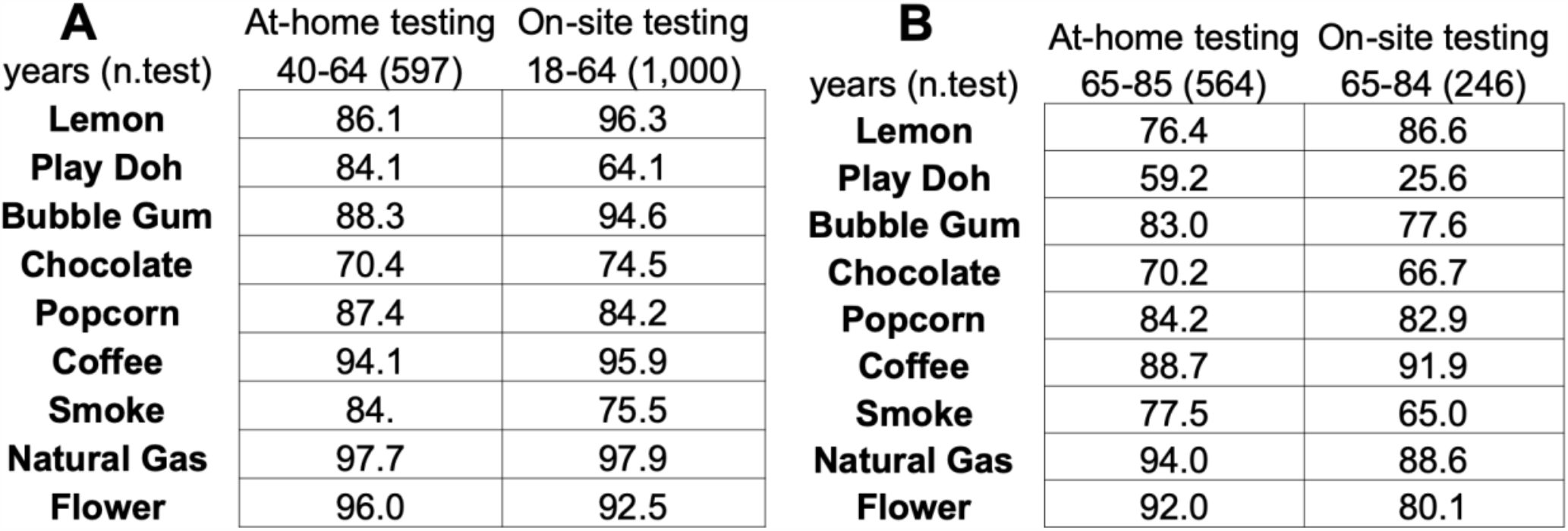
Comparison of correct responses obtained performing the test On-site (normative data) and At-home. A) Percentage of individuals younger than 65 that correctly identified the odorant. B) Percentage of individuals older than 65 that correctly identified the odorant. At-home testing results are similar compared to On-site normative data.

Self-administration of the test reliably detected the acknowledged gender-sensitivity effect, in which females are shown to perform with higher accuracy on an odor identification test. Accuracy was higher for women compared to men after controlling for participants’ age, F(1, 1160) = 22.953, *p* <.0001. The effect size calculated as Hedge’s g, was 0.41 and is very similar to the effect size reported in the literature. A recent metanalysis study using data from 106 papers, confirmed that women have a better sense of smell compared to males, [3].

Administration of this test remotely also replicated the age-related loss in olfactory acuity, participants were clustered into 5 decade-groups F(4, 1157) = 11.266, *p* <.0001, that begins around the fifth decade of life [2], consistent with what is observed when the test is administered in a testing facility (figure 3).

**Figure 3.**
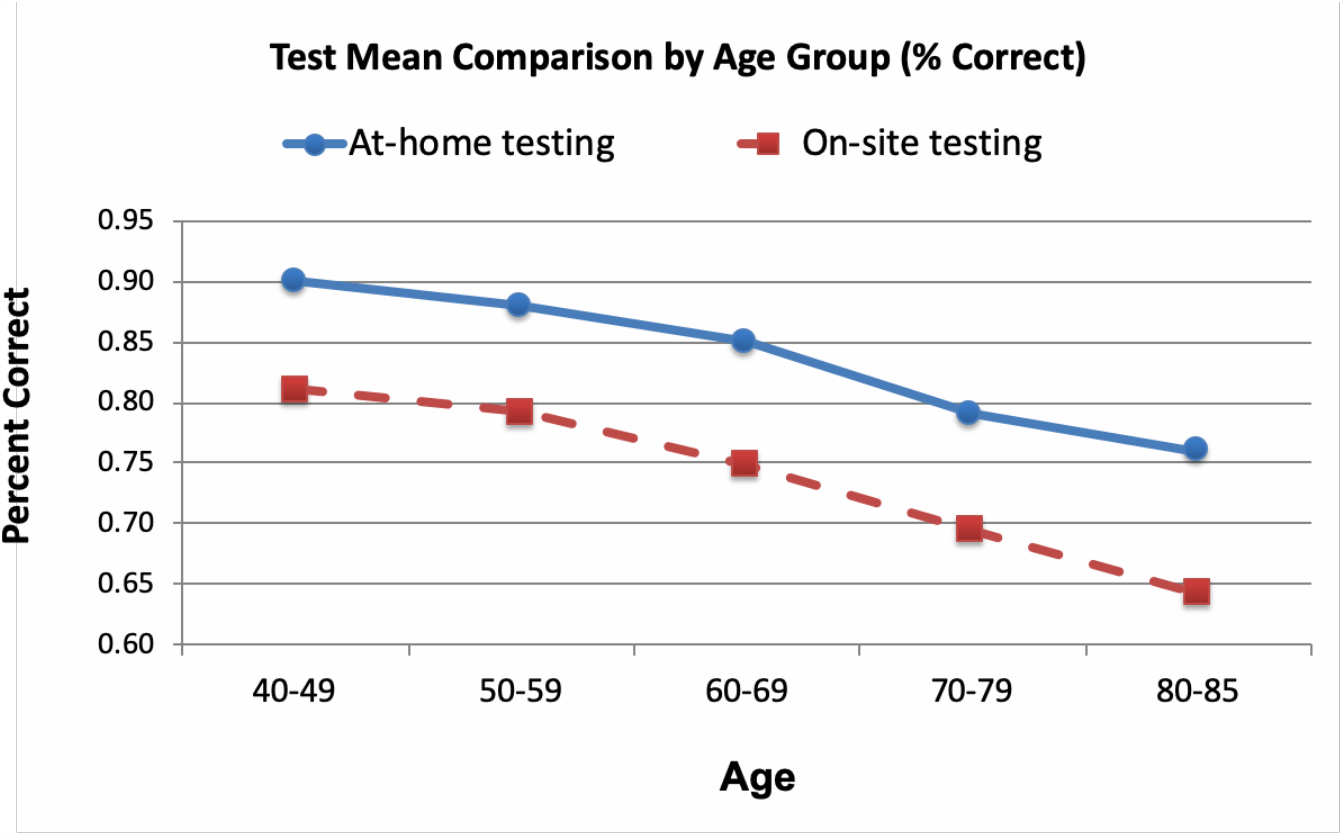
Validation of the NIH toolbox test performed At-home vs On-site. Mean proportion correct score for the At-home test (solid line) compared to the On-site test (dashed line). Age-dependent olfactory decline can be detected using the NIH odor identification cards.

## Discussion / Conclusion

In this study we explored the possibility of using the NIH Toolbox Odor ID Test as an inexpensive and convenient method to test olfactory performance remotely. From recent studies, a picture has emerged indicating that measuring olfaction has widespread utility since it seems to be a very good indicator of overall health and a potential correlate of cognitive dysfunction.

Based on this study we found that the NIH Toolbox Odor ID Test can be self-administered at home with comparable results to normative data obtained in a research setting. Further, remote use of the NIH Toolbox Odor ID Test is sensitive enough to detect age-dependent olfactory decline and gender-dependent sensitivity. In short, this feasibility study indicates that using the NIH Toolbox Odor ID Test is an appropriate method to measure olfactory performance via self-administration.

### Study funding

This research was funded, in part, under a 2015 Formula Grant with the Pennsylvania Department of Health. The Department specifically disclaims responsibility for any analyses, interpretations or conclusions.

## Data Availability

Data may be accessed upon request from the authorship team

## Author’s contributions

study concept and design PD, CJ, drafting the manuscript CJ, data analysis CJ, PD, CM, data interpretation and revising the manuscript CJ, CM, SM, MWW, DT, RN, MC, PD.

